# A Meta-Analysis of Mental Health among Latino Adults: Elucidating Disparities from Differences

**DOI:** 10.1101/2021.06.04.21255741

**Authors:** Karen R. Flórez, Kyra Alyssa C. Abbu, Fariha Hossain, Aprielle Wills, Joshua Breslau

**Author notes:** **Corresponding Author**: Karen R. Flórez, DrPH, MPH, CUNY Graduate School of Public Health and Health Policy, 55 West 125^th^ Street, New York NY 10027 USA.

## Abstract

Latinos continue to experience disparities in access to treatment for mental health, and these appear to be worsening with time. This meta-analysis identified and compared studies (N=20): (1) Across Latino origin groups, (2) Across immigration-related characteristics, and (3) with non-Latino groups. For the first comparison, results consistently showed Puerto Ricans had the highest rate of utilization compared to their Mexican, Cuban, Central American, and Other counterparts. For the second comparison, U.S-born Latinos had higher utilization and mental health costs for services, as well as higher rate of depression/anxiety symptomatology compared to their immigrant counterparts. For the third comparison, results were not as consistent but trended towards lower/less rates by Latinos compared to non-Latino whites. More research is needed on the different Latino groups and across the acculturation spectrum if we are to understand disparities from differences in risk and service use among this heterogeneous group.

## Introduction

Latinos continue to experience acute disparities in access to treatment for mental health, and these appear to be worsening with time. For example, in 2008, 57.4% of all Latino adults experiencing a major depressive episode received treatment in the last 12 months and in 2016 only 52.7% received care.^1^ In 2016, Latino adults (52.7%) were less likely than white adults (67.2%) to receive treatment for depression and this trend has persisted since the first National Healthcare Quality and Disparities Report was published in 2008.^1^ Other indicators show that Latino patients who receive treatment may not have their treatment needs met adequately. For example, Latinos are less likely to receive care from a physician that is culturally and linguistically congruent, and they rely more often on their primary care physician for mental health services.^2^ Primary care practices have been found to be less likely to use care management processes for a chronic illness like depression compared to chronic physical illnesses like asthma or diabetes, leading to a lack of adequate care for depression in the primary care setting.^3^

However, comparisons between Latinos and non-Latino Whites are of limited utility, because of the heterogeneity of the Latino population with respect to country of origin, migration history, and reception in the United States there are stark differences in disease risk.^4^ For instance, there is consistent evidence of differences across Latino groups defined by country of origin in the relationship between immigrant generation and risk for psychiatric disorder.^2^ Across multiple studies, there are no differences between island-born and mainland-born Puerto Ricans and large differences between immigrant and US-born Mexican-Americans.^5^ The extent to which evidence regarding disparities in mental health or mental health services use are consistent or variable across Latino subgroups is not known.

There are three main types of comparisons that researchers have used to understand disparities in mental health care affecting Latinos, each providing a different perspective on the same underlying issues. First, they have compared Latinos and specific Latino subgroups with non-Hispanic Whites. These comparisons are important for identifying disparities. Where Latinos are divided into sub-groups, these comparisons can help identify heterogeneity. Second, some studies make comparisons among Latino subgroups based on countries of origin. For example, Cubans and Mexicans. Third, some studies make comparisons within Latino groups based on characteristics related to immigration and acculturation. For example, years living in the United States. These different comparisons should produce a consistent body of evidence while taking into account the heterogeneity of this group. It is useful to focus on all three because the evidence based on each one is limited and the combination provides additional detail and replication, which is important for establishing robust results.

This paper reviews the recent literature on the heterogeneity of mental health disparities among Latinos. Our goal is to identify which sub-groups have been studied, where gaps remain in the empirical literature, and whether the existing literature has produced consistent or inconsistent results. We draw on the Institute of Medicine (IOM)’s conceptual framework.^6^ Specifically, this framework attempts to distinguish mere differences from disparities by focusing on factors above-and-beyond differences in prevalence, need, eligibility, and preferences. Instead, factors like discrimination, bias, stereotyping and the “ecology” of features within the medical system that enable disparities and inequity to develop.^6^ We were also particularly interested in the reference population that Latinos are being compared to, since this has major implications in how we conceptualize and address disparities among this group. We also sought to include all areas of mental health, which we categorize as (1) access, (2) participation, (3) nature and quality of services, and (4) mental health outcomes/symptoms.

## Methods

### Evidence acquisition

A two-prong search strategy was employed to identify journal articles. First, multiple databases were searched (i.e., PUBMED, Web of Science) using various combinations of search terms (e.g., ((behavioral health) OR (mental health) OR (substance use)) AND ((service use) OR (utilization) OR (health care)) AND ((Hispanic) OR (Latino) OR (Mexicans) OR (Mexican Americans) OR (Cubans) OR (Puerto Ricans)) AND (Disparity)) through October 2020. Publications were restricted to those written in English and conducted in the U.S. Second, bibliographies of the studies included in previous related literature reviews were reviewed to ensure a comprehensive approach.

Figure 1 shows our methodology of search and inclusion/exclusion. The search yielded 1,303 journal articles, of which 410 were removed for being duplicates or non-relevant material based on title review. Out of the 893 articles screened, 776 were excluded due to irrelevance, or because they were a thesis or book. From the 117 studies whose title and abstract were reviewed, we only included original studies that used empirical, quantitative data. After applying these criteria, the results were reduced to 21 relevant articles. However, one article was further excluded because it was reporting the effects of a specific type of program^7^ for a total of 20 articles.

**FIGURE 1.**
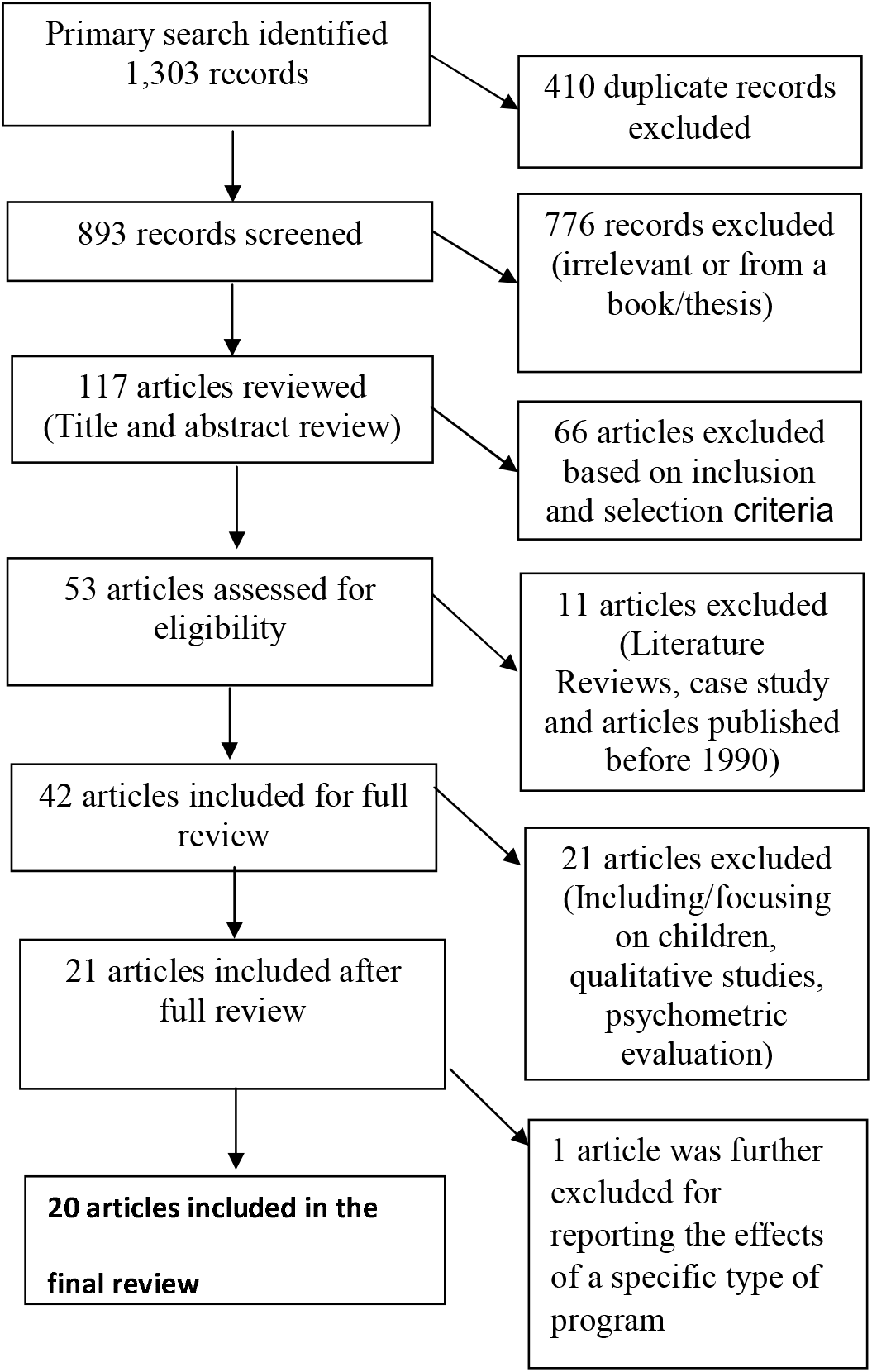
Flow diagram for literature search and review.

### Evidence synthesis

Methods for conducting literature reviews developed for the Effective Public Health Practice Project were used for data extraction using the Quality Assessment Tool for Quantitative Studies.^8^ This approach was used since it facilitated systematic evaluation of each study through a standardized form that classified and described study design, confounders, data collection details, and analyses. Two independent reviewers conducted the data abstraction and verified the results of the data points: type of comparison made (i.e., across Latino origin groups; across immigration-related characteristics; with U.S.-born non-Latino groups); sample; covariates; type of mental health outcome (i.e., mental health and functioning; service use; quality of care).

### Data analysis

Methods in creating forest plots for three comparisons used a step-by-step guide for calculations from BioMed Central^9^ and used RStudio^10^ for the data visualization. The measures of association for every article in the three comparison groups were determined. Since the calculations tool is applicable for articles that measure prevalence, articles that do not use prevalence are excluded from the forest plots. The first comparison is across Latino origin groups, in which articles included in the forest plot focused on Mexican, Puerto Rican, Cuban, Central American, and Other.^11-16^ Only one article was exclude from the forest plot because the comparison was between Puerto Ricans in the mainland U.S compared to those in the island.^17^ The second comparison is across immigration-related characteristics,^12,15,16,18-21^ and three articles were excluded from the forest plot given their operationalization of immigration-related characteristics.^16,18,22^ Here, the reference group is U.S. born and the other years are aggregated to calculate the prevalence for foreign-born. The third comparison is across non-Latino groups,^23-27^ and some were excluded from the forest plot as well.^28-34^ Additional details specific to some articles are noted in each figure.

## Results

Table 1 provides descriptive information of studies in the sample (N=20), organized by the type of comparison being made. Articles are listed more than once if they report more than one type of comparison. Specifically, comparison groups are in the shaded rows, and include (1) across Latino-origin groups, (2) across immigration-related characteristics (e.g., nativity) and (3) with U.S.-born non-Latino origin groups (e.g., non-Latino whites). In the first broad category of studies focused on the comparison across major Latino origin groups except for Canino that focused on Puerto Ricans.^17^ Most studies analyzed data from large nationally representative samples, mainly from the National Latino and Asian American Studies (NLAAS). Two solely focused on women;^11,13^ all other studies adjusted for gender, but varied substantially in the set of socioeconomic covariates used in adjusted models and how each was modeled. For example, poverty was sometimes a dichotomous variable reflecting whether the person was above or below the federal poverty level or income levels. Level of educational attainment was not modeled consistently, nor did most of the studies mention how educational level equivalency was reached given the different structures between the U.S. system relative to Latin America. Employment status was not consistently controlled for, and marital status as well as insurance coverage were sometimes treated as proxy for SES.

**Table 1:**
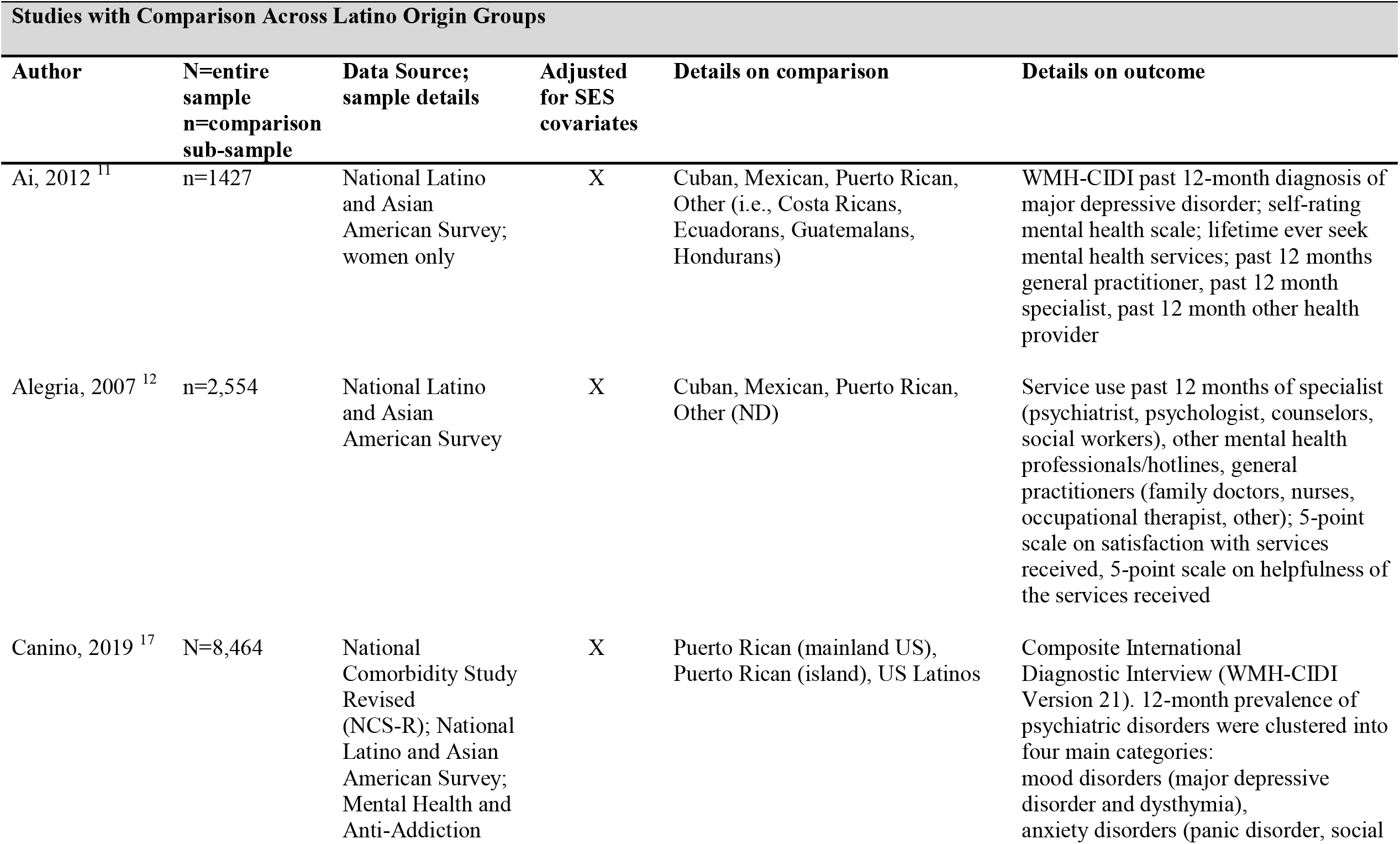
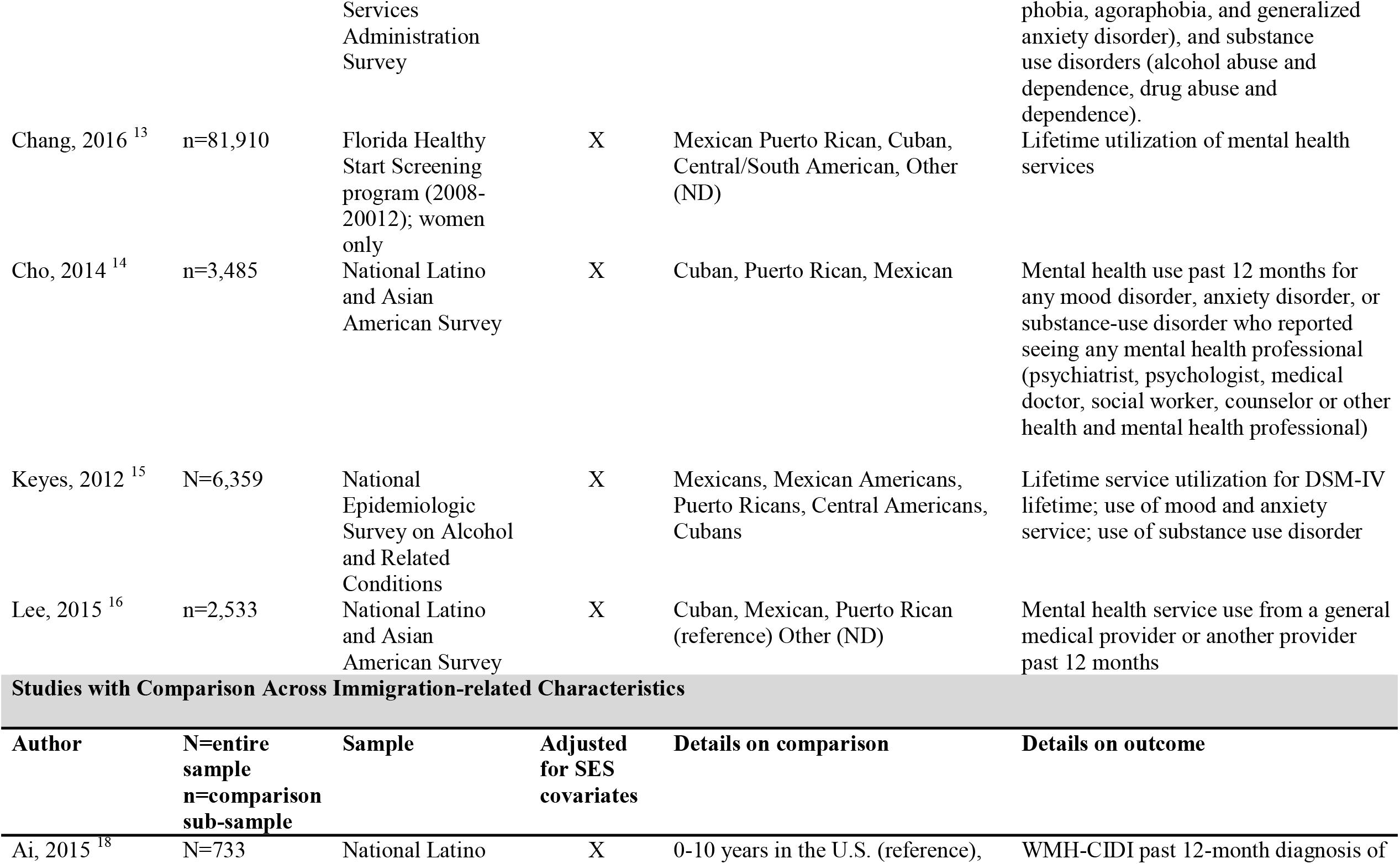
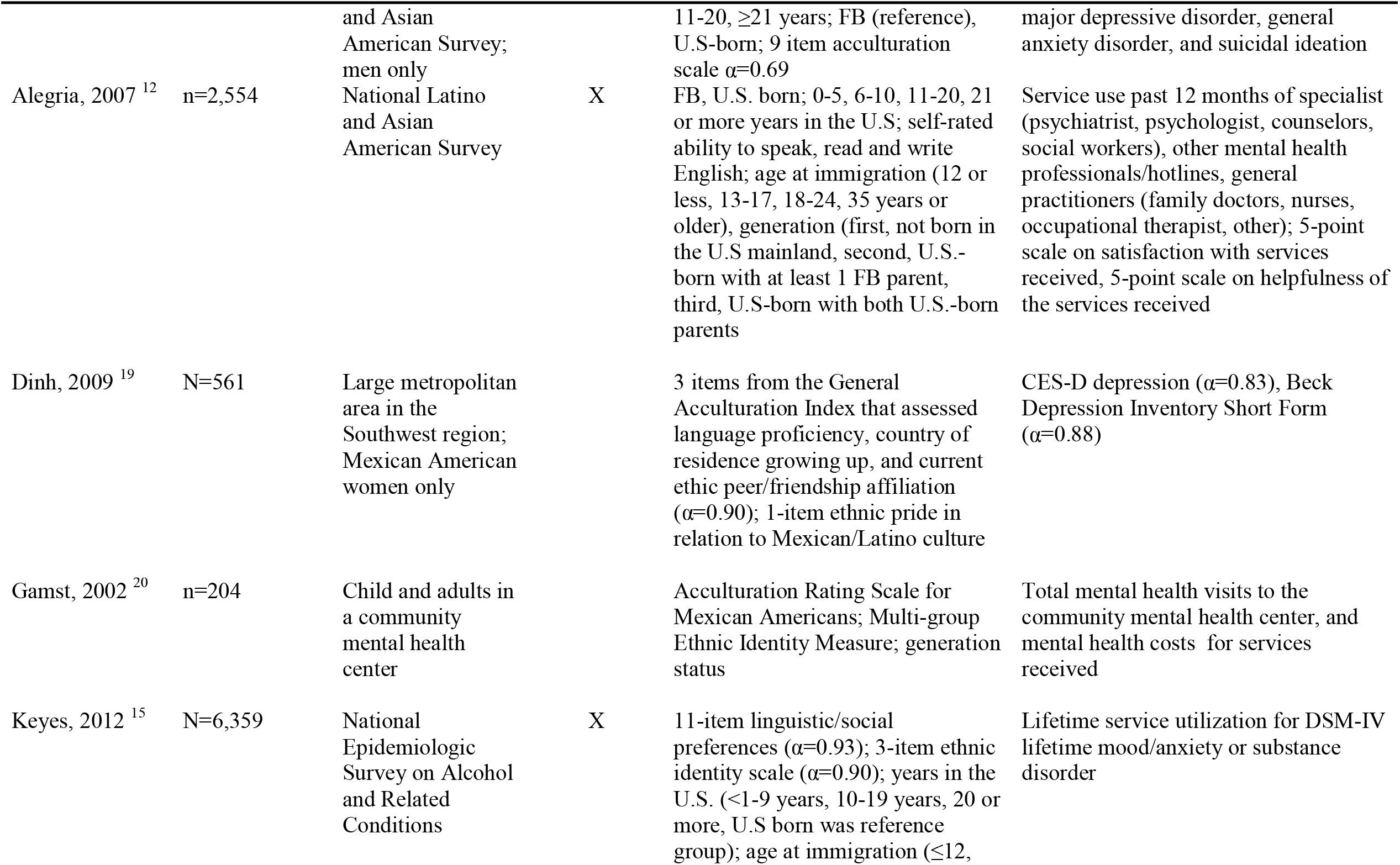
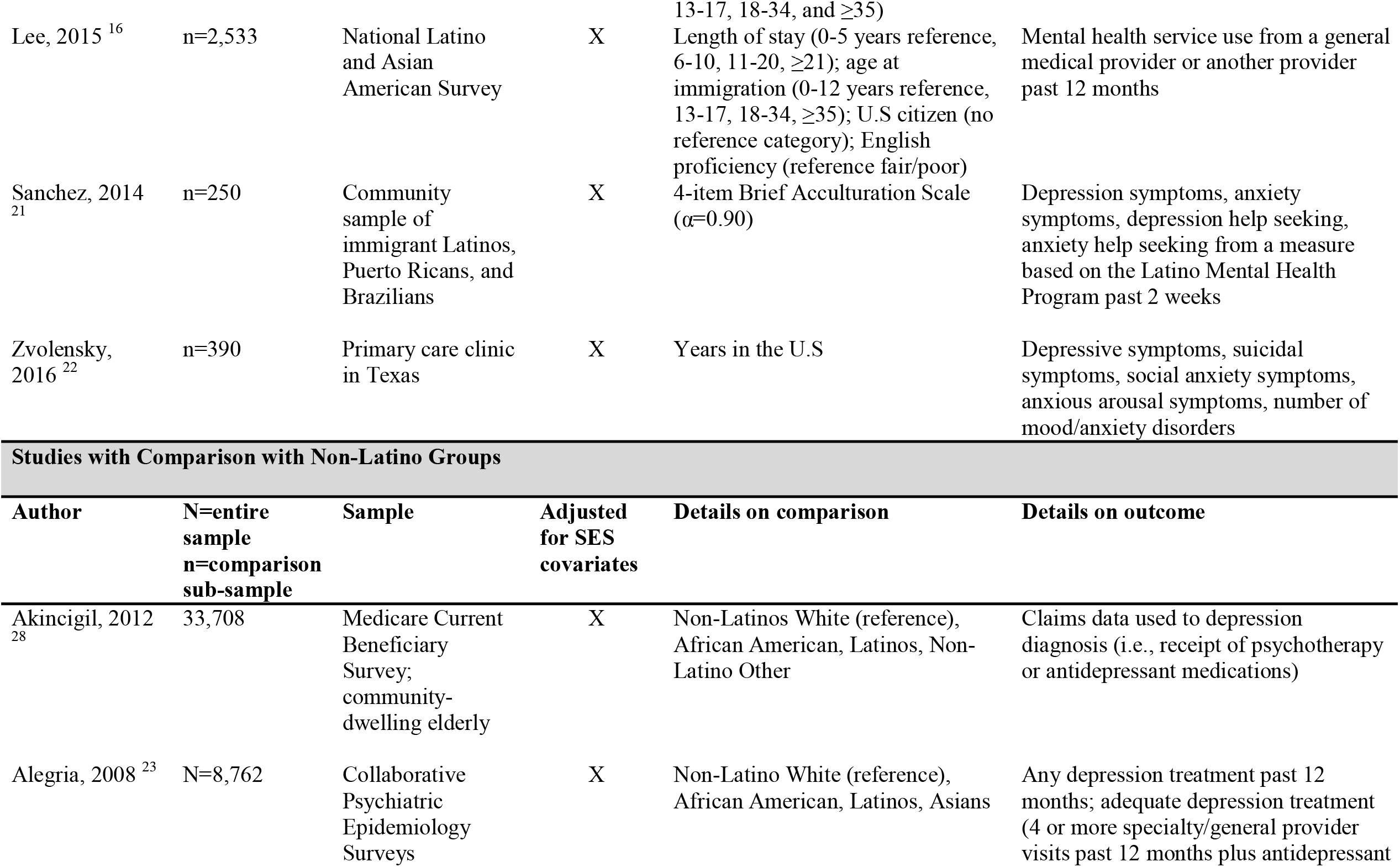
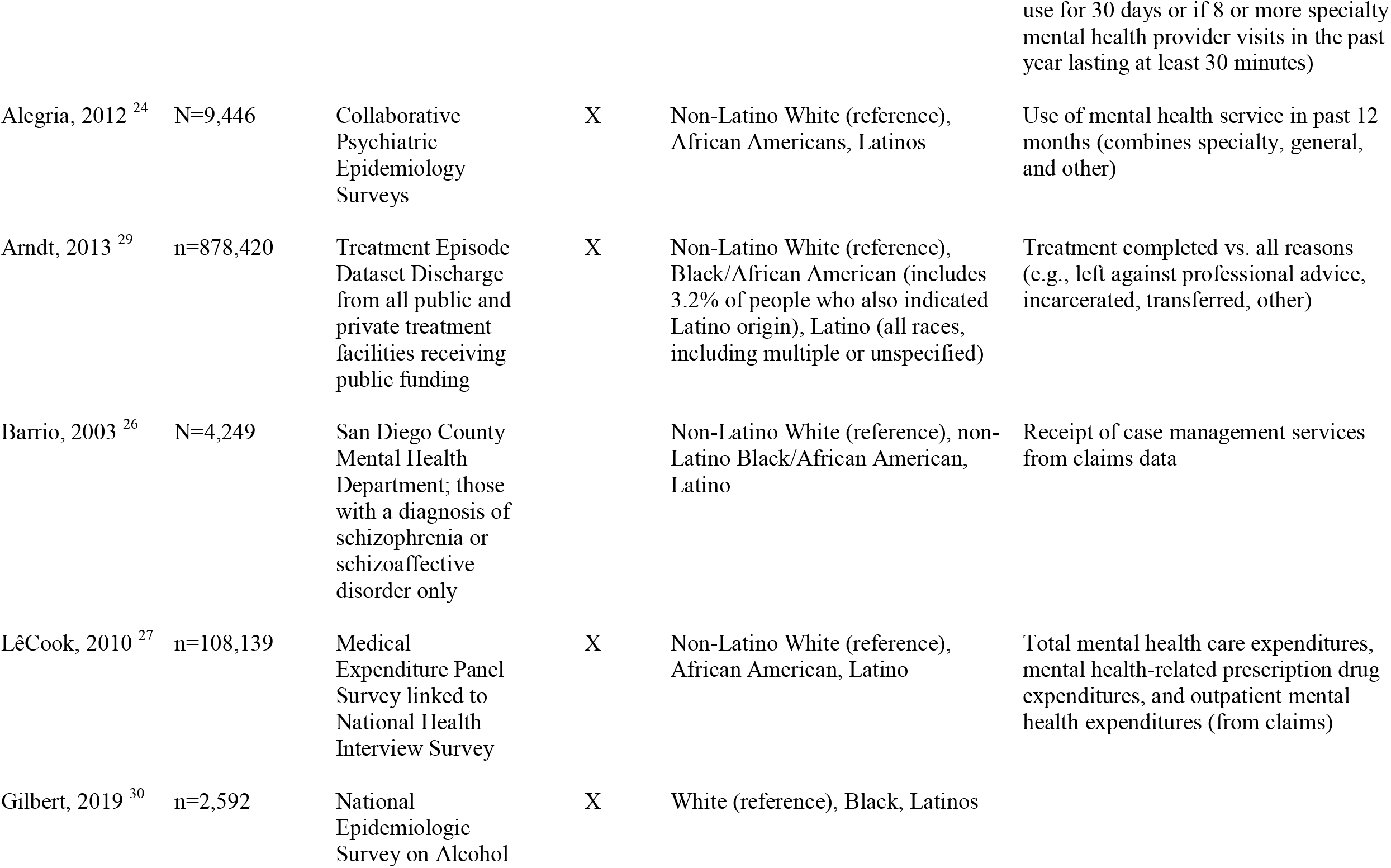
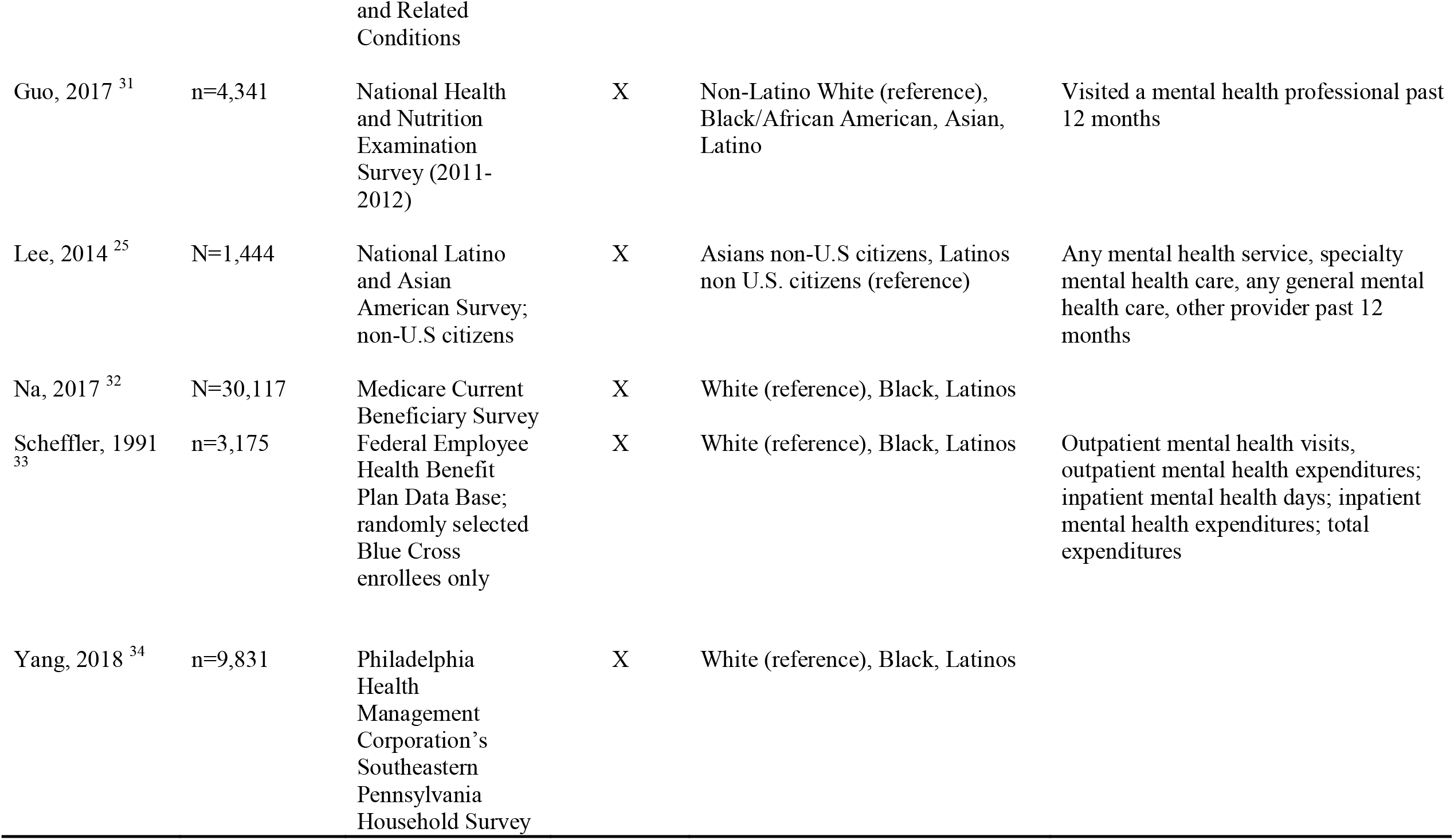
Descriptive information of studies in the sample (N=20), by comparison

Most studies included the major Latino subgroups of Puerto Ricans, Mexicans, as well as an “other” category.^11-13,16^ However, most studies did not define the group except for Ai, 2012, who was explicit about combining Costa Ricans, Ecuadorians, Guatemalans, and Hondurans.^11^ Keyes et al., was also the only study to separate Mexicans from Mexican Americans based on self-report;^15^ whereas Cho et al., dropped those that were not Cuban, Puerto Rican, or Mexican from the analysis.^14^ There was variation in the way in which statistical analyses were performed and/or presented.^11-13^ Only one study use Mexicans as the reference category in the logistic regression models, but results were only presented for Puerto Ricans and not Cubans.^14^ Puerto Ricans were used as the reference group in the other two studies although no rationale for this decision was presented.^15,16^ Outcomes for this set of studies were mostly focused on utilization;^12-16^ though there was some variation on whether the utilization was over the lifetime ^13,15^ or in the past 12 months.^11,12,14,16^ Only one study included mental health outcomes like self-rated mental health and diagnosis of major depression^11^ and only one study included quality of care outcomes.^12^

Table 1 also provides descriptive information for the second broad category of studies that focused on the comparison across immigration-related characteristics. These sets of studies had more variation regarding data scope; some used large nationally representative samples ^12,15,16,18^ whereas others relied on convenience samples.^19-22^ There was substantial variation across studies in the way in which the exposure was measured. Three studies did not use any acculturation scales but relied solely on proxy-measures like nativity (foreign-born vs. U.S. born);^12^ age at immigration (0-12, 13-17, 18-34, ≥35)^12,16^, self-rated ability to speak, read, and write English that was then collapsed into excellent/good vs. fair/poor by Lee et al., 2015 or into predominant language of Spanish, bilingual, and English by Alegria et al., 2007. Length of stay in U.S. also varied across studies with some using a categorical variable (0-5, 6-10, 11-20, ≥21)^12,16^ or modeled as continuous variable of years lived in the U.S.^22^ Generation status^12^ and citizenship status^16^ were not used as much. Some studies used a combination of the more widely used proxy measures like length of stay in the U.S. and nativity along with acculturation or ethnic affiliation scales,^15,18^ whereas other studies simply relied on scales to measure this construct.^19-21^ Most studies reporting psychometric properties of scales exhibited good to excellent reliability, except for one study that reported a Cronbach’s α=0.69.^18^ Once again, there was substantial variation in the way in which statistical analyses were performed and/or presented, even in the studies using the same dataset and/or variables.^12,15,16,18^ This set of studies focused equally on mental health symptoms and/or functioning ^18,19,21,22^ and mental health service use.^12,15,16,20^ Only one also included quality of care outcomes.^12^

Lastly, table 1 depicts descriptive information for the third category of studies that focused on the comparison of Latinos with U.S.-born non-Latino group. All studies used large datasets included in the other sets of studies (e.g., Collaborative Psychiatric Epidemiology Surveys, which includes NLAAS),^23-25^ but some used unique data sources like Southeastern Pennsylvania Household Survey,^34^ National Epidemiologic Survey on Alcohol and Related Conditions,^30^ Medicare Current Beneficiary Survey,^28,32^ National Treatment Episode Discharge Dataset,^29^ Medical Expenditure Panel Survey linked to NHIS,^27^ NHANES,^31^ and the Federal Employee Health Benefit Plan Database.^33^ One study focused solely on a region in California; San Diego County.^26^ A few of the studies also focused on specific sub-populations like those with a diagnosis of schizophrenia or schizoaffective disorder,^26^ U.S. citizens vs. non-citizens,^25^ and Blue Cross enrollees.^33^ Only the latter randomly selected this subsample; however, this resulted in the Latino cell being too small for accurate calculation.^33^ These studies did consistently choose Whites as the reference category,^23,24,26-29,31,33^ and most were explicit in their methods in only including non-Latino Whites except for Scheffler et al.^33^ Yet, studies were not as explicit in the treatment of Latinos identifying as Black/African American, nor was Black/African American used as reference category in any of the studies. Only one study made the comparison between Latinos and Asians.^25^ Mental health use was again the focus of most studies; however, some studies derived this outcome from claims data ^26-29,33^ rather than self-report use.^23-25,31^

There was also more variation in the way in which service mental health use was operationalized, which included expenditures,^27,33^ or treatment completion or receipt of case management services.^26,29^ Only one included adequacy of treatment as a measure of quality of care ^23^ and mental health symptom.^28^

Forest Plot 1 compares mental health service and prevalence across Latino origin groups, specifically between Mexican, Puerto Rican, Cuban, Central American, and Other categories. The summary measure reflects the overall results from all the studies in Forest Plot 1. It shows that Puerto Ricans have the highest rate of lifetime utilization of mental health services, as well as utilization in the past 12 months compared to their Mexican, Cuban, Central American, and Other counterparts.

Forest Plot 2 compares studies using immigration-related characteristics; specifically, U.S-born and foreign-born Latinos. It excludes four studies from Table 1 grouped with comparison across immigration-related characteristics due for not measuring prevalence. Under this category, there are four outcomes containing one study each. Across all four outcomes, U.S-born Latinos had a higher mental health service use in the past 12 months through a specialist, higher total mental health costs for services, higher lifetime utilization of mental health services, and higher rate of depression/anxiety symptomatology. The summary score across all categories shows this consistency in results across studies.

Forest Plot 3 compares Latinos with non-Latino groups. Under this group, there are two outcomes. The first outcome is the mental health service use in the past 12 months. Alegria, 2008 ^23^ and Lee, 2014 ^25^ measured non-Latino White and, in both studies, they have used these services the highest compared to other groups. However, Alegria, 2008 and Lee, 2014 measured non-Latino Asian, which has lower mental health service use than Latinos.^23,25^ The results were less consistent when the comparison was with Asian Americans and non-Latino Black/African Americans. Results were also not as consistent for total mental health expenditure, though the summary score does show lower/less rates by Latinos compared to non-Latino whites across this range of mental health outcomes.

## Discussion

This literature review found that the disparities documented by Latino adults regarding access to mental health care varies depending on the reference group used. This variation is important since the landmark disparities framework urges research to distinguish disparities from mere differences. ^6^ This review identified 3 major comparisons: those made across Latino subgroups; those across immigration-related characteristics; those made between Latinos and a non-Latino group. Within each of the comparisons, however, there were remarkable consistency.

Specifically, in the comparison across Latino subgroups, Puerto Ricans had the highest rate of mental health service use across all studies. One the one hand, research has historically highlighted the various socioeconomic vulnerabilities experienced by Puerto Ricans (e.g., high unemployment rate, poverty, living in high-crime segregated areas),^35^ which may place them at greater risk for psychopathology, and thus, greater need for mental health use. However, the correlation between lower SES and increased psychopathology does not always hold.^17^ Puerto Ricans may also have greater access to healthcare related resources, including insurance relative to other Latino subgroups,^36^ which may fuel their mental health service usage.

There was also consistency in the comparison across immigration-related characteristics. Specifically, the studies show less/lower rates among immigrants; those with less time in the U.S; and Spanish proficiency. Taken together, these studies are in line with other studies showing an association between dwindling physical health profiles with greater acculturation. However, this linear process, with often zero-sum assumptions has been extensively critiqued, ^37,38^ as well the assumption that the acculturating group is homogenous (i.e., that the group represents a common cohesive culture).^39^ This is faulty particularly in the study of Latinos as stark differences in cultural models and sociopolitical trajectories have been established in this group,^4^ and which in turn, may be associated with different psychopathologies. For example, subgroups exposed to severe social and political violence (e.g., Guatemalans, Colombians, Salvadorians) may be particularly susceptible certain psychopathologies (e.g., post-traumatic-stress-disorder as oppose to depressive symptomology) and little research has focused on the intergenerational transmission of this trauma from immigrants to their U.S.-born children.

Lastly, there was consistency in the studies comparing Latinos with other non-Latino groups in that the former had significantly lower/less rates compared to non-Latino whites across a range of mental health outcomes. In general, Latinos are treated in the literature as a group with robust social networks that may be able to buffer harmful health affects even in the face of poverty and other SES-related conditions. This could include mental health outcomes. However, others point to the increasingly dehumanizing anti-Latino rhetoric of the US environment, irrespective of immigration status.^40^ This form of psychological violence, coupled with pressures at school, work, or with family and peers, may create an untenable environment that fosters anxiety, depression, and other psychopathologies. Further, structural racism in the healthcare setting may further exacerbate any mental health needs among Latinos, especially among Afro-Latinos or those that phenotypically cannot pass as white.^41^

Taken together, this review highlights some important patterns in mental health disparities among Latinos. It reviews more recent literature on the subject and uses the Institute of Medicine’s framework to attempt to distinguish mere differences from disparities.^6^ We also sought to include all areas of mental health, and moved beyond Andersen’s model of predisposing, enabling, and need factors that does not contextualize the experience of accessing health care by vulnerable communities within their lived realities. However, the review also has some limitations given the lack of research that uses comparable acculturation variables across Latino groups and with similar mental health outcomes.

Despite these limitations, this review does contribute to the field by parsing out some of the heterogeneity of the Latino population with respect to country of origin, migration history, and reception in the United State to further understand patterns in this important and growing population. To truly move the field forward; however, more longitudinal research is needed on the different Latino subgroups and across the acculturation spectrum to detect whether convergence occurs over generations in the U.S. in terms of mental health risk and service use. This will also be crucial at distinguishing disparities from mere differences and embolden efforts to tackle more systemic factors like discrimination, bias and stereotyping within the mental health care system.

## Data Availability

Not applicable

## Acknowledgements

This paper was sponsored and supported by the National Institute of Minority Health and Health Disparities under award numbers: R01MD010274; PI: Breslau and R01MD010274-04S1; PI: Flórez. No potential conflicts of interest were reported. The findings and conclusions in this report are those of the authors and do not necessarily represent the official position of the funding organization.

---

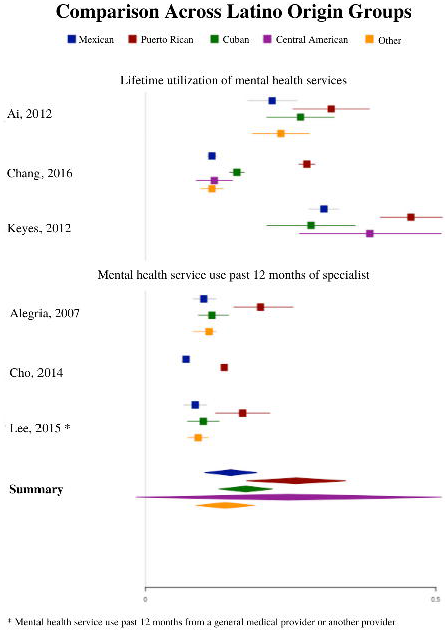

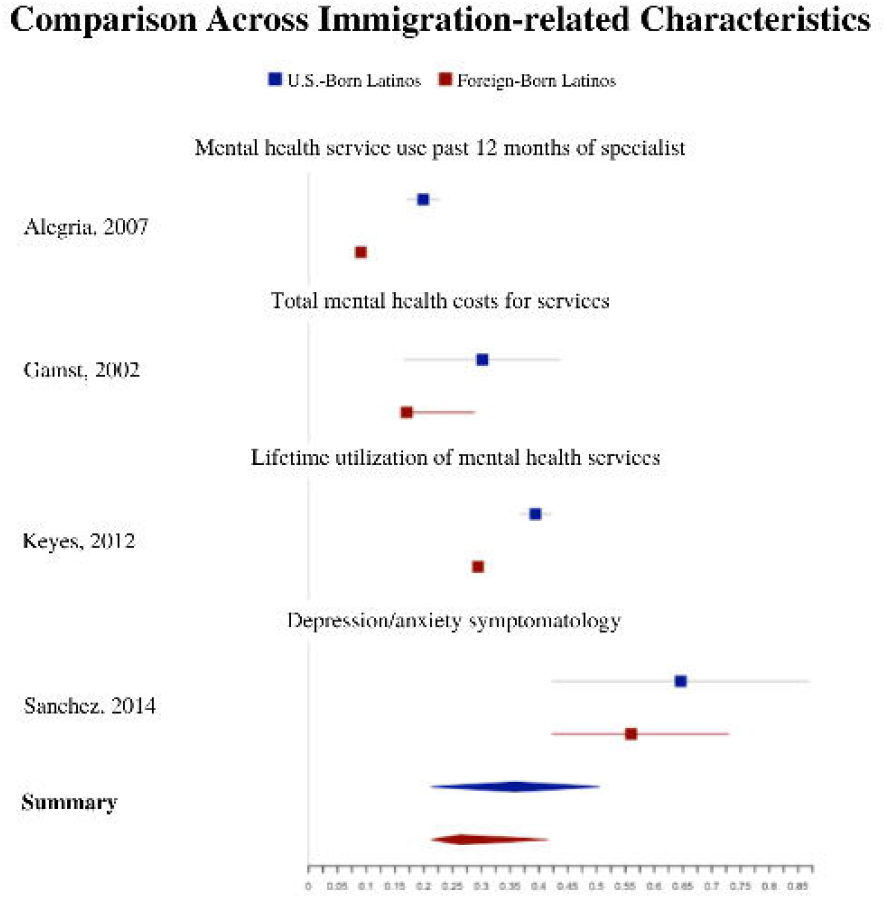

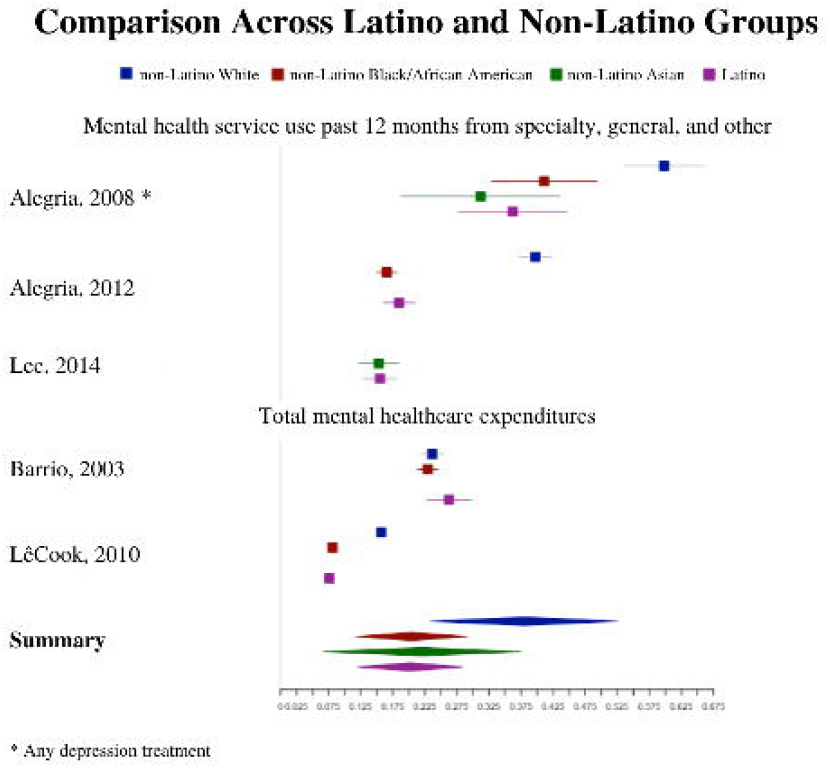

